# Endovascular thrombectomy for acute ischemic stroke with a large infarct area: an updated systematic review and meta-analysis of randomized controlled trials

**DOI:** 10.1101/2024.04.02.24305181

**Authors:** Shivani Ravipati, Ayesha Amjad, Komal Zulfiqar, Hannah Biju, Wajeeh Hassan, Haider Mumtaz Jafri, Ali Husnain, Ibrahim Tahir, Muaaz Aslam, Sharib Afzal, Muhammad Ehsan, Huzaifa Ahmad Cheema, Muhammad Ayyan, Wajeeh Ur Rehman, Sourbha S. Dani

## Abstract

**Background:** Since the efficacy and safety of endovascular thrombectomy (EVT) in patients with acute ischemic stroke with a large infarct area is still inconclusive, we sought to compare functional and neurological outcomes with the use of endovascular thrombectomy versus medical care alone.

**Methods:** We searched MEDLINE (via PubMed), Embase, Cochrane Library, ClinicalTrials.gov, and the International Clinical Trials Registry Platform (ICTRP) to retrieve all the relevant randomized controlled trials (RCTs) on this topic. Review manager (RevMan) was used to perform meta-analyses using a random-effect model. Dichotomous outcomes were pooled using risk ratios (RR) with 95% confidence intervals (CIs).

**Results:** Our meta-analysis included 6 RCTs with a total of 1665 patients. Most studies included patients with an ASPECTS score of 3-5. Our results demonstrate that endovascular thrombectomy significantly increased the rates of functional independence (mRS ≤ 2) (RR, 2.49; 95% CI, 1.89-3.29) and moderate neurological outcome (mRS ≤ 3) (RR, 1.90; 95% CI, 1.50-2.40) at 90 days. The benefit of EVT for these outcomes remained the same at 1-year follow-up.. Endovascular thrombectomy was associated with increased rates of early neurological improvement (RR, 2.22; 95% CI, 1.53-3.22), excellent neurological recovery (mRS ≤ 1) (RR, 1.75; 95% CI, 1.02-3.03), and decreased rate of poor neurological recovery (mRS 4-6) (RR, 0.81; 95% CI, 0.76-0.86). No significant difference was found between the two groups regarding all-cause mortality (RR, 0.86; 95% CI, 0.72-1.02), decompressive craniectomy (RR, 1.32; 95% CI, 0.89-1.94), and the incidence of serious adverse effects (RR, 1.39; 95% CI, 0.83-2.32) between the two groups. Endovascular thrombectomy significantly increased the rates of any intracranial hemorrhage (RR, 1.94; 95% CI, 1.48-2.53) and symptomatic intracranial hemorrhage (RR, 1.73; 95% CI, 1.11-2.69).

**Conclusion:** Endovascular thrombectomy (EVT) significantly improves neurological and functional outcomes in patients with acute ischemic stroke with a large infarct area (ASPECTS 3-5) compared to medical therapy alone, with an increased risk of symptomatic intracranial hemorrhage.

## Introduction

Over the last decade, studies have showcased a clear benefit of endovascular thrombectomy (EVT) over medical therapy (MEDT) alone in managing acute ischemic stroke (AIS) due to large vessel occlusion (LVO) (1). Therefore, current guidelines advocate for EVT in individuals presenting with Alberta Stroke Program Early CT Score (ASPECTS) ≥6 (range: 0-10, inverse relation with the infarction size) (2,3). However, patients with large-core infarct (LCI) – defined as ischemic volume greater than 50 mL or 70 mL with cerebral perfusion <30%, or ASPECTS <6 – were not considered candidates for EVT due to the potential risk of reperfusion injury, hemorrhagic conversion, and perception of negligible advantage as a notable portion of brain tissue being already infarcted in these patients (4). The management of this patient population, constituting one-fourth of LVO cases, revolved mainly around curtailing secondary insult from raised intracranial pressure and cerebral edema in the background of large ischemic volume. Frequently applied approaches included but not limited to optimizing cerebral blood flow and diligent blood pressure monitoring, bolstering collateral circulation in at-risk tissue, seizure identification, and management, and preventing recurrent recurrence (4,5). In brief, the LCI stroke population remained underrepresented for a long time in studies assessing the benefits of EVT over MEDT.

RESCUE-Japan LIMIT trial by Yoshimura et al. was the first randomized controlled trial (RCT) to evaluate the efficacy and safety of EVT in this subset population (6). The trial reported favorable outcomes in the EVT arm compared to MEDT alone and paved the way for further studies. As a result, five more RCTs have been conducted to date (5,7–10). Several meta-analyses meticulously combine the results of published trials in the literature (1,11–13). However, these reviews,do not include all trials as results from some trials, such as LASTE, were reported after publication. They do not include all trials as results from some trials, such as LASTE, were reported after publication (10). Furthermore, we also incorporated the 90-day results from the TESLA and SELECT2 trials, which helped assess the long-term efficacy of EVT (14,15).

This updated meta-analysis aims to comprehensively synthesize the latest results from all trials, offering higher-level insight into the efficacy and safety of EVT and MEDT vs. MEDT alone.

## Materials and methods

This systematic review and meta-analysis followed the guidelines outlined in the Cochrane Handbook for Systematic Reviews (16) and reported according to the Preferred Reporting Items for Systematic Reviews and Meta-Analysis (PRISMA) statement (17). Ethical approval was not required for this study. The protocol was registered with the International Prospective Register of Systematic Reviews (PROSPERO) under the identifier CRD42023492739.

### Data Sources and Search Strategy

Electronic searches were conducted using various online databases from inception to December 2023. These included the Cochrane Central Register of Controlled Trials (CENTRAL), MEDLINE (via PubMed), Embase (via Ovid), ClinicalTrials.gov, and the International Clinical Trials Registry Platform (ICTRP). No geographical or language restrictions were applied during the search process. Additionally, the reference lists of the included studies and similar systematic reviews were screened to retrieve relevant articles. The detailed search strategy for MEDLINE is given in Supplementary Table 1.

### Eligibility Criteria

The inclusion criteria for the study were as follows: (1) study design: randomized controlled trials (RCT); (2) population: individuals with large ischemic stroke, defined as a large vessel occlusion with an ASPECTS score of 3 to 5; (3) intervention: endovascular thrombectomy plus medical therapy; (4) control group: medical therapy alone; and (5) outcome: reporting at least 1 outcome of interest.

The exclusion criteria were as follows: (1) study designs other than RCTs, such as quasi-randomized trials, reviews, and observational studies; (2) studies conducted on animals; and (3) studies evaluating outcomes in patients who have undergone ischemic stroke with a small infarct area.

### Study Selection

The literature search results were imported into Zotero, a software management tool for screening articles. After de-duplication, two authors independently conducted the initial phase of screening titles and abstracts. The full-text screening was performed on the remaining studies, and a final selection was made based on adherence to our eligibility criteria. A third author settled any disagreements regarding the selection of the studies. The selection process is presented in the form of a PRISMA flow chart.

### Data collection process and data items

Two review authors extracted data from the included studies into a pre-piloted structured Excel spreadsheet. Relevant data items were extracted, including study characteristics (country, study design, total participants, intervention, main inclusion criteria, ASPECTS score, study follow-up duration, and baseline imaging), baseline characteristics (age, sex, number of patients in each group, NIHSS score, infarct core volume, ASPECTS score, and occlusion location), primary outcomes (functional Independence (mRS ≤ 2) and moderate neurological outcome (mRS ≤ 3)), and secondary outcomes (all-cause mortality at 90 days, early neurological improvement, excellent neurological recovery (mRS ≤ 1), poor neurological recovery (mRS ≤ 4-6), any intracranial hemorrhage, symptomatic intracranial hemorrhage, decompressive craniectomy, and >1 SAE (serious adverse effect).

### Risk of Bias and Quality Assessment

The risk of bias was assessed in the included studies using the revised Cochrane Risk of Bias Tool for randomized controlled trials (RoB 2.0) (18). The domains that were evaluated included (1) risk of bias resulting from the randomization process, (2) risk of bias due to deviation from the intended intervention, (3) risk of bias due to missing outcome data, (4) risk of bias in measuring the outcome, (5) risk of bias in selecting the reported results, and (6) other bias. For clarification, “low” indicates a low risk of bias, and “high” indicates a high risk of bias. If the study lacked information or had uncertainty over the potential for bias, the item was judged as “unclear.” Any disagreements in evaluating the risk of bias were resolved through discussion to reach a consensus between the two authors. A third author served as an arbiter if needed.

### Statistical Analysis

Statistical analyses were performed using the Review Manager software (version 5.3; RevMan v5.3). The DerSimonian-Laird variance estimator was used to apply a random effects model. Dichotomous outcomes were pooled using risk ratios (RR) with 95% confidence intervals (CIs). Heterogeneity was assessed using the chi-square and I-square tests. The alpha level for the chi-square test was set at 0.1, as determined by the Cochrane Handbook, and was considered to suggest statistically significant heterogeneity.

## Results

### Search Results and Study Selection

A total of 2077 studies were identified from various databases. Following deduplication and initial screening, 55 full-length articles were assessed for eligibility. A total of 6 RCTs were included in our systematic review and meta-analysis. The detailed screening process is illustrated in the PRISMA flowchart (Figure 1)

**Figure 1.**
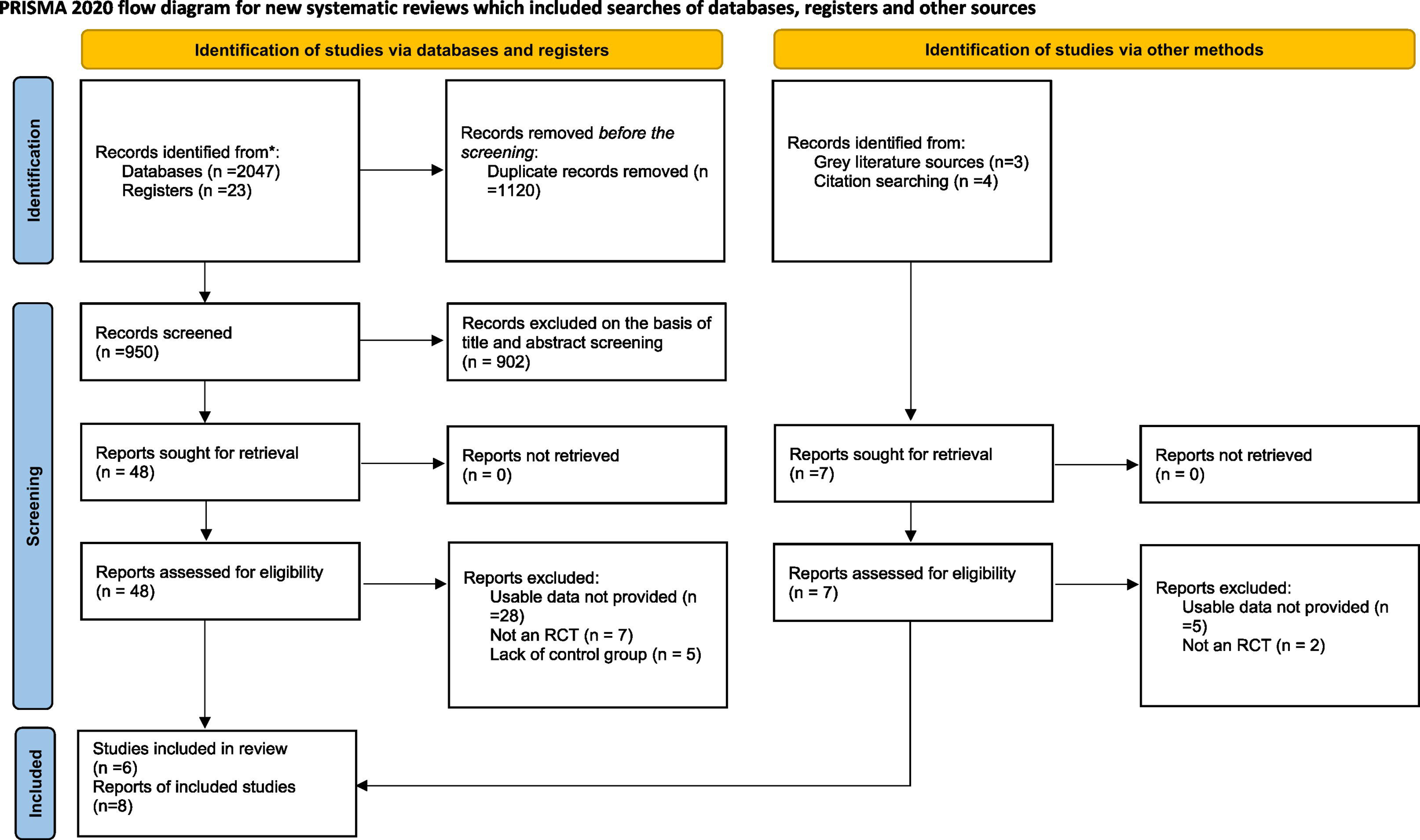
PRISMA 2020 flow chart of included and excluded trials. PRISMA, Preferred Reporting Items for Systematic Reviews and Meta-Analyses.

### Characteristics of Included Studies

We included 6 RCTs in our meta-analysis (5–10). A total of 1665 patients were included, of which 945 received endovascular thrombectomy with medical therapy, and 942 received medical therapy alone. All studies except LASTE included patients having a pre-stroke mRS score of 0–1 and a stroke with a large ischemic-core volume, defined as an ASPECT score of 3 to 5. The follow-up duration was 90 days in all the studies. Reports of two studies reporting outcomes at 1-year follow-up have also been included in this systematic review. Detailed characteristics of included studies are given in Table 1

**Table 1.**
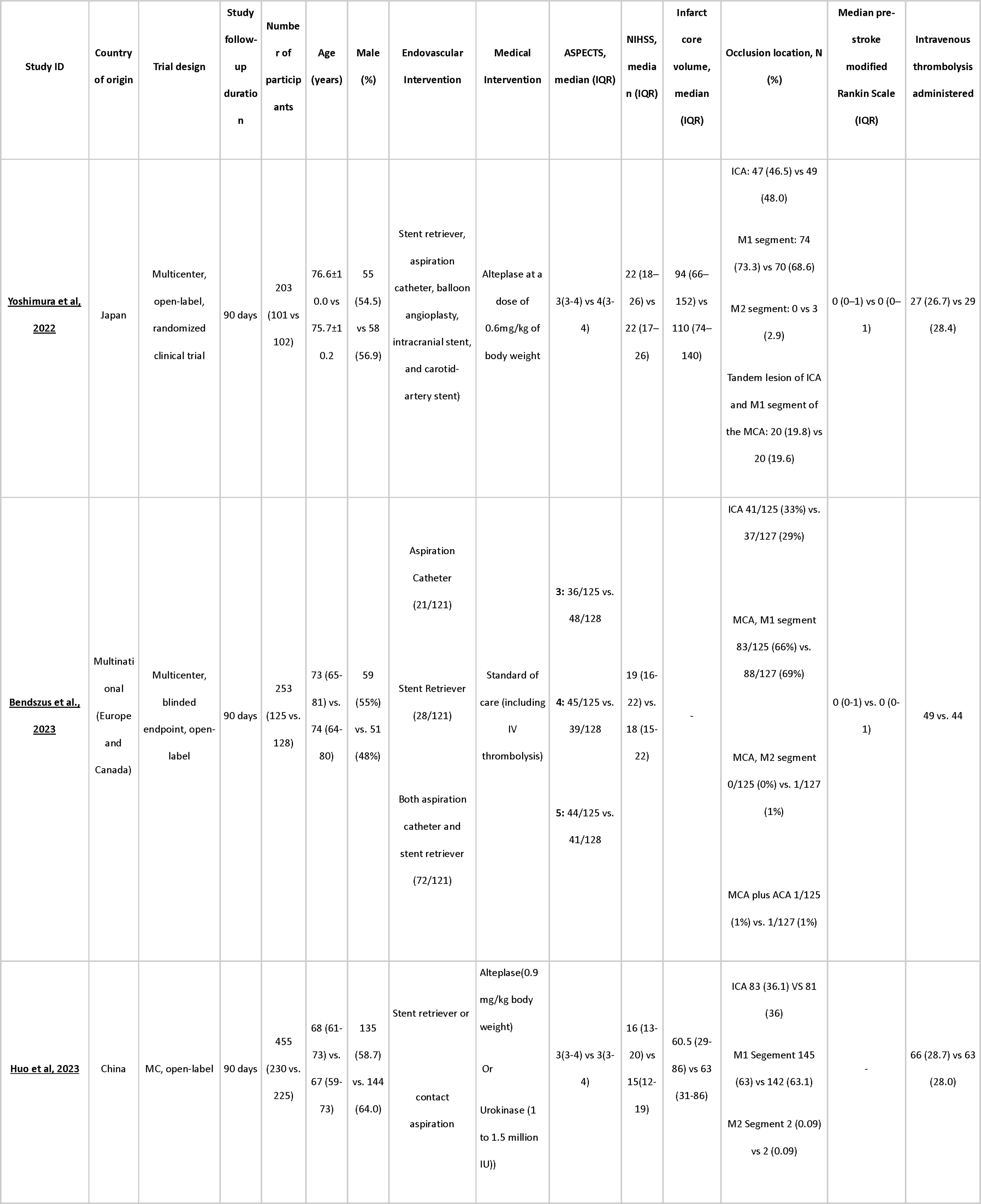
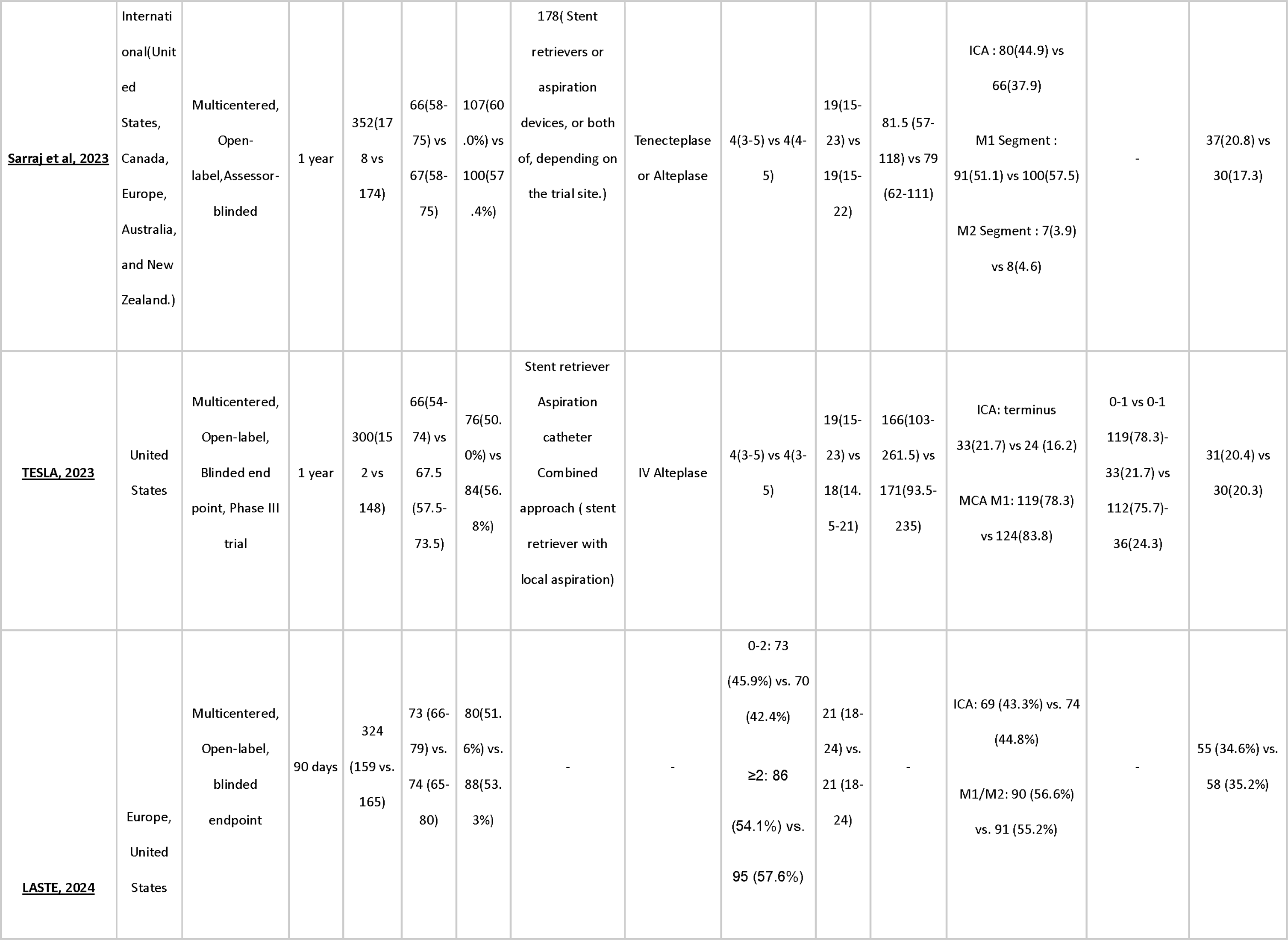
Study characteristics of included studies.

### Risk of Bias and Quality of Evidence

We used Cochrane RoB2 to evaluate the risk of bias of included studies. All six RCTs were deemed to have a low risk of bias [Figure 2].

**Figure 2.**
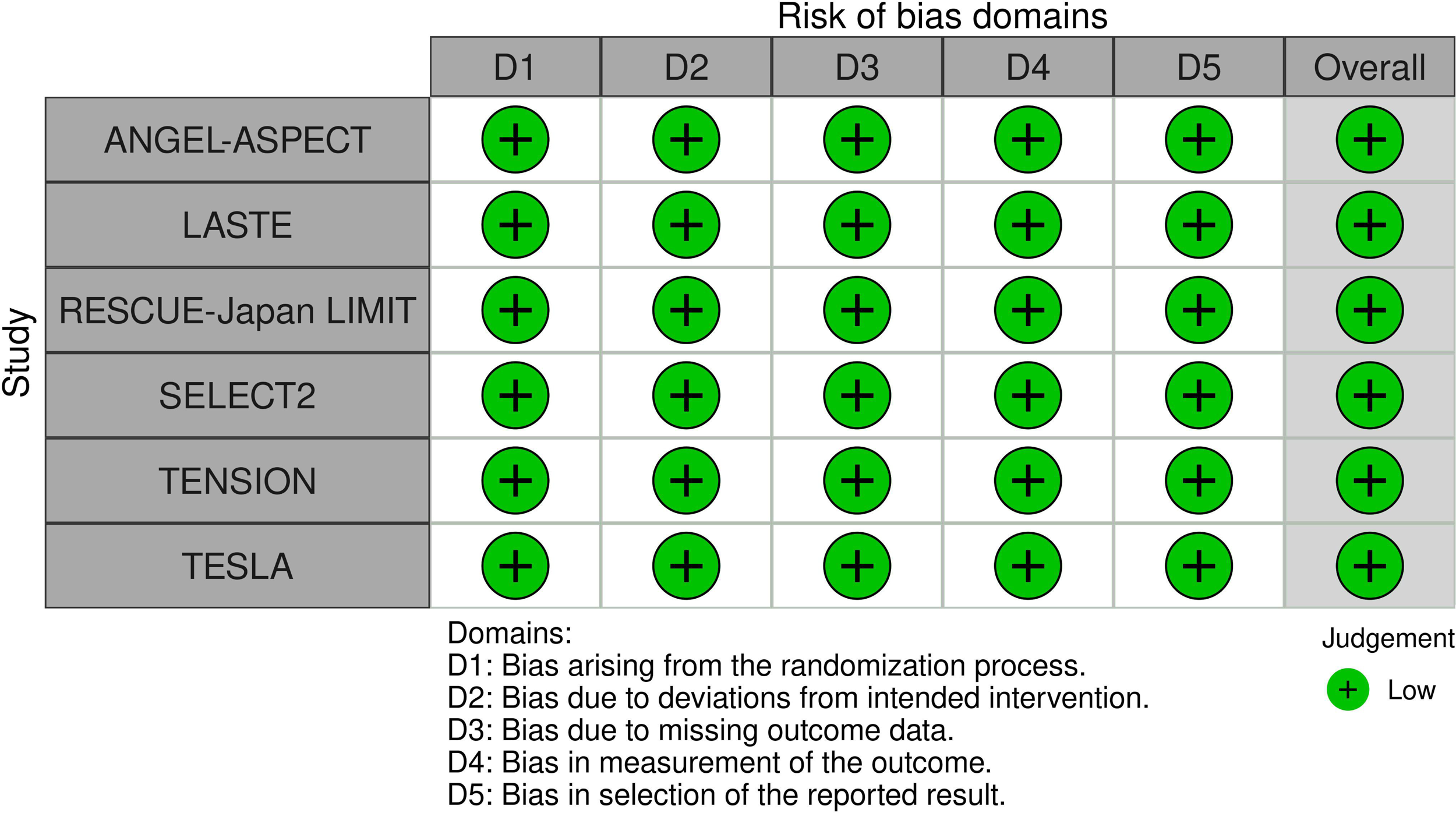
Quality assessment of randomized controlled trials (RCTs).

## Primary Outcomes

### Functional Independence (mRS ≤ 2)

Our meta-analysis of 6 trials indicates that endovascular thrombectomy significantly increased the rate of functional Independence (mRS ≤ 2) at 90 days (RR, 2.49; 95% CI, 1.89-3.29; Figure 3). The heterogeneity reported between studies for this outcome was low (I^2^ =7%). A meta-analysis of only two trials found endovascular thrombectomy to significantly increase the rate of functional Independence (mRS ≤ 2) at 1 year (RR, 3.84; 95% CI, 2.35 – 6.29; Supplementary Figure 1) with minimal heterogeneity (I^2^ = 0%)

**Figure 3.**
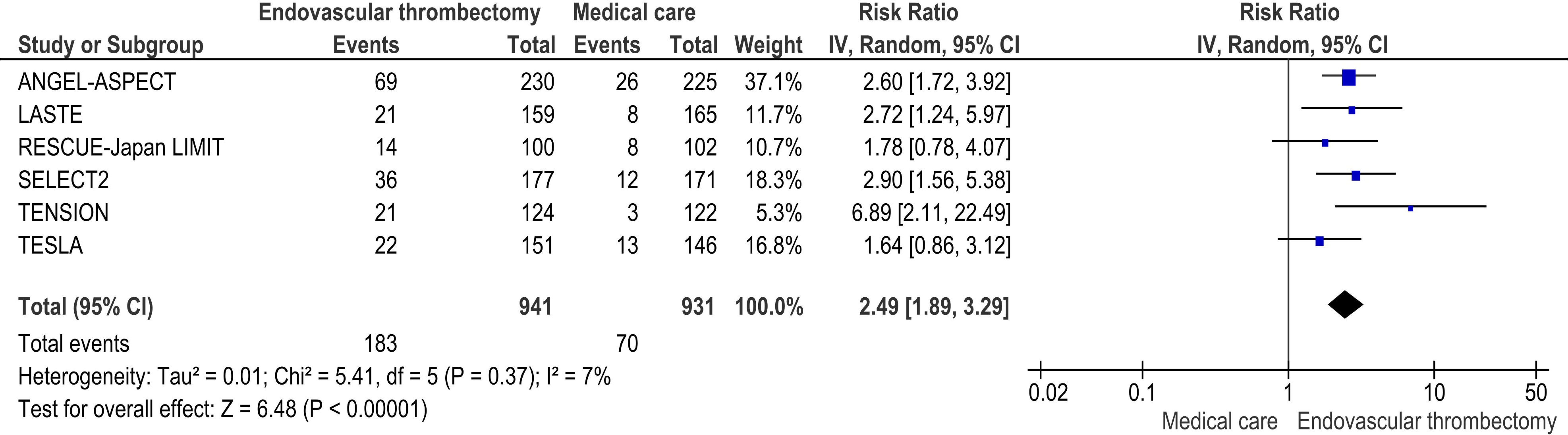
Forest plot of functional independence (mRS ≤ 2) at 90 days follow-up.

### Moderate Neurological Outcome (mRS ≤ 3)

Endovascular thrombectomy significantly increased the rate of independent ambulation (also known as moderate neurological outcome) compared to medical therapy (RR, 1.90; 95% CI, 1.50-2.40; Figure 4). Statistical heterogeneity was found to be moderate (I^2^ = 51%).

**Figure 4.**
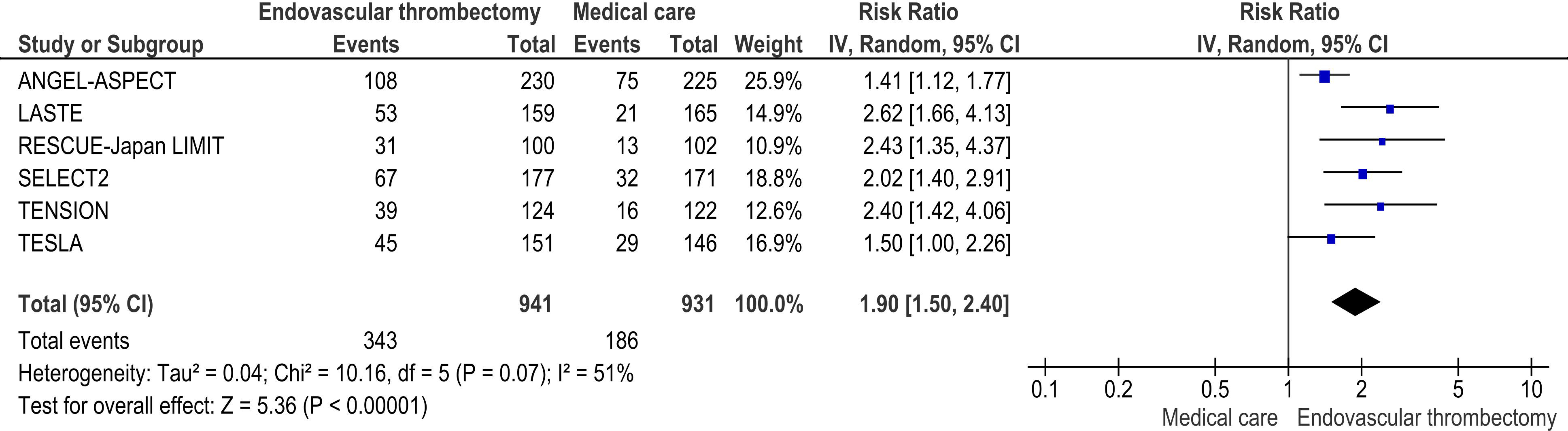
Forest plot of independent ambulation (mRS ≤ 3) at 90 days follow-up.

A meta-analysis of only two trials found endovascular thrombectomy to significantly increase the rate of independent ambulation (mRS ≤ 3) at 1 year (RR, 2.08; 95% CI, 1.56 – 2.77; Supplementary Figure 2) with minimal heterogeneity (I^2^ = 0%).

## Secondary Outcomes

### Early Neurological Improvement

Our meta-analysis found that endovascular thrombectomy increased the rate of early neurological improvement (RR, 2.22; 95% CI, 1.53-3.22; Supplementary Figure 3) with minimal statistical heterogeneity (I^2^ = 17%).

### Excellent Neurological recovery (mRS ≤ 1)

Endovascular thrombectomy was associated with an improved rate of excellent neurological recovery (mRS ≤ 1) as compared to medical therapy alone (RR, 1.75; 95% CI, 1.02-3.03; Supplementary Figure 4). The inter-study statistical heterogeneity was estimated to be low (I^2^ = 36%).

### Poor Neurological Recovery (mRS 4-6)

Endovascular thrombectomy was associated with a significantly decreased rate of poor neurological recovery (mRS 4-6) as compared to medical therapy alone (RR, 0.81; 95% CI, 0.76-0.86; Supplementary Figure 5) with minimal statistical heterogeneity (I^2^ = 0%).

### All-cause Mortality

No statistically significant difference was found between the two groups regarding all-cause mortality (RR, 0.86; 95% CI, 0.72-1.02; Supplementary Figure 6) with moderate statistical heterogeneity (I^2^ = 45%).

### Symptomatic Intracranial Hemorrhage

Endovascular thrombectomy significantly increased the rate of symptomatic intracranial hemorrhage (RR, 1.73; 95% CI, 1.11-2.69; Supplementary Figure 7). Statistical heterogeneity was found to be minimal (I^2^ = 0%).

### Decompressive craniectomy

No statistically significant difference was found between the two groups regarding decompressive craniectomy (RR, 1.32; 95% CI, 0.89-1.94; Supplementary Figure 8). Statistical heterogeneity between studies was low (I^2^ = 41%).

### Any intracranial hemorrhage

Endovascular thrombectomy significantly increased the rate of any intracranial hemorrhage (RR, 1.94; 95% CI, 1.48-2.53; Supplementary Figure 9). The estimated heterogeneity was substantial (I^2^ = 72%).

### >1 SAE

Our meta-analysis showed no significant difference in rates of serious adverse effects between the two groups (RR, 1.39; 95% CI, 0.83-2.32; Supplementary Figure 10) with substantial heterogeneity (I^2^ = 93%)

## Discussion

Our meta-analysis included six randomized controlled trials (RCTs), which showed that endovascular thrombectomy (EVT) in combination with medical therapy significantly improves outcomes for patients suffering from large vessel occlusion strokes. EVT increases the likelihood of primary outcomes, i.e., functional independence and moderate neurological outcomes at 90 days, with maintenance of these benefits at 1-year follow-up. Moreover, our analysis indicates that EVT leads to higher rates of early neurological improvement and excellent neurological recovery while reducing the incidence of poor neurological recovery. Despite these benefits, the procedure is associated with an increased risk of symptomatic intracranial hemorrhage; however, it does not significantly affect all-cause mortality or the need for a decompressive craniotomy.

AHA/ASA recommends EVT for patients with an ASPECT score greater than 5. In contrast, those with ASPECT scores below 5 have a poor prognosis and are at risk of symptomatic intracranial hemorrhage (19). Our findings align with the HERMES meta-analysis, suggesting that a lower ASPECT score could still warrant consideration, especially in younger patients (20). There was considerable heterogeneity regarding inclusion criteria among the six trials. The trials in our study had diverse ASPECT scores and ischemic core volumes as their criteria for inclusion, with the majority having ASPECT 3-5. The LASTE trial (10) also included patients with ASPECT scores ranging from 0 to 2. The trials SELECT-2 (5) and ANGEL-ASPECT (7) have incorporated ischemic regions quantified in volumes determined using imaging with more than 50 ml and 70–100 ml, respectively. Our analysis indicated that EVT enhances functional outcomes in post-stroke patients across a wide range of stroke severities, mRS scores, and ischemic volumes.

Our results are consistent with the previous two meta-analyses (21,22), except for the incidence of symptomatic intracranial hemorrhage (sICH); our analysis revealed an increased risk of sICH. This difference can be attributed to including two latest trials, TESLA(8) and LASTE (10). The LASTE trial carried significant weight in the meta-analysis and indicated a heightened risk of sICH, likely accounting for this deviation from earlier reviews.

Considerable heterogeneity was observed in the time elapsed from stroke onset to intervention across all trials. Most trials have set 24 hours as the maximum time for intervention from the time of symptom onset. Notably, the TENSION trial included patients within 12 hours of stroke onset, while the RESCUE Japan LIMIT and LASTE trials included patients who presented within 6 hours of stroke onset. The onset-to-intervention time window is crucial for the patients because the odds of independent ambulation and discharge to home are reduced by 8% and 10% for every 60 minutes of onset-to-intervention time extended in the early window (within 6 hours). In the late window (6–24 hours), the chances of independent ambulation and discharge to home are 1% and 2% lower for every 60-minute delay in treatment.

Hence, early treatment is more favorable for patient recovery and is associated with lower 90-day mortality (23).

ASPECT scores determine the areas with ischemia but do not precisely measure infarction volume or extent. While diagnostic imaging like CT and MRI perfusion can determine the volume of infarcted tissue, it cannot ultimately determine the actual tissue viability. Nevertheless, EVT reperfusion of the ischemic area can be beneficial. The increased blood supply could alleviate edema and remove toxins, leading to neurological recovery regardless of the ischemia area/volume (24,25). Further studies or imaging techniques are required to understand the pathophysiological changes following reperfusion.

The primary strength of our meta-analysis lies in the comprehensive selection of the latest randomized controlled trials (RCTs) and the meticulous adherence to appropriate inclusion and exclusion criteria. Furthermore, we also incorporated the 90-day results from the TESLA and SELECT2 trials (14,15) for our primary outcomes, aiding in the evaluation of the long-term effects of EVT. The inclusion of the latest RCTs with large sample sizes increased the statistical power of our meta-analysis.

However, there are a few limitations to the articles that we included. Firstly, all the studies were of an open-label design, which may introduce bias due to lack of blinding, potentially influencing the outcomes through placebo effects or altered reporting symptoms. Secondly, early termination of studies or trials might have overestimated the results as premature conclusions drawn from incomplete data may inflate the perceived benefits of the treatment. Third, regional specificities observed in studies conducted exclusively in Japan and China, such as different standard doses of rt-PA and varying prevalences of intracranial artery stenosis, respectively, could limit our findings’ generalizability to a global population. Additionally, exclusion criteria for patients older than or equal to 80 years used by Huo et al. (7) might affect generalizability since it excluded a significant portion of the stroke population from the results.

In addition, two studies focusing on a specific imaging technique (non-contrast CT) raised concerns about the generalizability of their results to settings where other imaging methods are commonly used (8,14). Furthermore, patient enrollment criteria based on the ASPECTS score led to the exclusion of patients with a score higher than 5 and with an infarct core volume of 70–100 ml in ANGEL-ASPECT And SELECT2 trials, resulting in potential bias in patient selection that may affect treatment outcomes. Additionally, Yoshimura et al. failed to document the causes of deaths during the study, raising questions about safety and associated risks related to the intervention (6).

Limitations in the review process include a small sample size, which might have reduced statistical power, reducing the ability to detect real differences between groups or variables being studied, resulting in false negative results and an increased risk of random error influencing the analysis. Additionally, there is moderate heterogeneity across certain aspects of our analysis, stemming from variations in study designs, patient populations, treatment protocols, or outcome measures among the included trials. This variation complicates the interpretation of specific sub-populations. Furthermore, sub-group analyses could not be performed based on patient age, stroke severity, or the presence of comorbid conditions, without which our conclusion may overlook significant variations in treatment efficacy. Lastly, individual patient data analysis was not performed due to insufficient data.

Future research should aim to standardize inclusion criteria, particularly regarding ASPECT scores and ischemic core volumes, to enhance and improve comparability among trials and robustness of meta-analysis. Investigating the impact of different inclusion criteria on treatment outcomes could provide valuable insights into patient selection. Long-term follow-up data from trials such as TESLA and SELECT-2 contribute to understanding EVT’s sustained benefits. Further studies with extended follow-up periods could elucidate EVT’s durability and identify potential late complications or recurrence risks. Future trials should prioritize blinding to minimize possible bias introduced by open-label designs. Additionally, efforts to prevent premature study termination and ensure complete data collection are essential to avoid overestimating treatment effects. Studies should strive for diverse patient populations and global representation to enhance the generalizability of findings. Addressing regional specificities, such as variations in treatment protocols and patient demographics, can facilitate more comprehensive recommendations applicable to diverse healthcare settings.

Overall, while the current evidence supports the efficacy of EVT in ischemic stroke with large infarct areas, ongoing research is essential to optimize patient selection, treatment protocols, and long-term management strategies, ensuring equitable access to effective interventions globally.

## Conclusion

Our meta-analysis demonstrates that endovascular thrombectomy (EVT) significantly improves neurological and functional outcomes in patients with acute ischemic stroke with a large infarct area (ASPECTS 3-5) compared to medical therapy alone. However, our findings also indicate a higher risk of symptomatic intracranial hemorrhage associated with EVT. Further studies are needed to evaluate the efficacy of EVT in patients with low ASPECTS scores (1–3) and confirm the safety profile of EVT.

## Data Availability

All data produced in the present study are available upon reasonable request to the authors

## Statements and Declarations

### Ethical Publication Statement

We confirm that we have read the Journal’s position on issues involved in ethical publication and affirm that this report is consistent with those guidelines.

### Declarations of interest

The authors declare that they have no conflicts of interest and no financial interests related to the material of this manuscript.

### Funding

No financial support was received for this study.

## Acknowledgments

N/A

## Data Availability Statement

Data will be provided on reasonable request from the corresponding author.

## Notes

### Competing Interest Statement

The authors have declared no competing interest.

### Funding Statement

This study did not receive any funding

## References

1. Kobeissi H, Adusumilli G, Ghozy S, Kadirvel R, Brinjikji W, Albers GW, et al. Endovascular thrombectomy for ischemic stroke with large core volume: An updated, post-TESLA systematic review and meta-analysis of the randomized trials. Interv Neuroradiol J Peritherapeutic Neuroradiol Surg Proced Relat Neurosci. 2023 Jun 28;15910199231185738.

2. Powers WJ, Rabinstein AA, Ackerson T, Adeoye OM, Bambakidis NC, Becker K, et al. Guidelines for the Early Management of Patients With Acute Ischemic Stroke: 2019 Update to the 2018 Guidelines for the Early Management of Acute Ischemic Stroke: A Guideline for Healthcare Professionals From the American Heart Association/American Stroke Association. Stroke. 2019 Dec;50(12):e344–418.

3. Turc G, Bhogal P, Fischer U, Khatri P, Lobotesis K, Mazighi M, et al. European Stroke Organisation (ESO) - European Society for Minimally Invasive Neurological Therapy (ESMINT) Guidelines on Mechanical Thrombectomy in Acute Ischemic Stroke. J Neurointerventional Surg. 2023 Aug;15(8):e8.

4. Migdady I, Johnson-Black PH, Leslie-Mazwi T, Malhotra R. Current and Emerging Endovascular and Neurocritical Care Management Strategies in Large-Core Ischemic Stroke. J Clin Med. 2023 Oct 20;12(20):6641.

5. Sarraj A, Hassan AE, Abraham MG, Ortega-Gutierrez S, Kasner SE, Hussain MS, et al. Trial of Endovascular Thrombectomy for Large Ischemic Strokes. N Engl J Med. 2023 Apr 6;388(14):1259–71.

6. Yoshimura S, Sakai N, Yamagami H, Uchida K, Beppu M, Toyoda K, et al. Endovascular Therapy for Acute Stroke with a Large Ischemic Region. N Engl J Med. 2022 Apr 7;386(14):1303–13.

7. Huo X, Ma G, Tong X, Zhang X, Pan Y, Nguyen TN, et al. Trial of Endovascular Therapy for Acute Ischemic Stroke with Large Infarct. N Engl J Med. 2023 Apr 6;388(14):1272– 83.

8. Intraarterial Treatment Versus No Intraarterial Treatment within 24 Hours in Patients with Ischaemic Stroke and Large Infarct on Noncontrast CT (TESLA): A Multicentre, Open-Label, Blinded-Endpoint, Randomised, Controlled, Phase 3 Trial by Albert J. Yoo, Osama O. Zaidat, Sami Al Kasab, Sunil A. Sheth, Ansaar T. Rai, Santiago Ortega-Gutierrez, Curtis A. Given, Syed F. Zaidi, Ramesh Grandhi, Hugo Cuellar, Maxim Mokin, Jeffrey M. Katz, Amer Alshekhlee, Muhammad A. Taqi, Sameer A. Ansari, Adnan H. Siddiqui, Nobl Barazangi, Joey D. English, Alberto Maud, Jawad Kirmani, Rishi Gupta, Dileep Yavagal, Jason Tarpley, Dhruvil J. Pandya, Marshall C. Cress, Sushrut Dharmadhikari, Kaiz Asif, Tareq Kass-Hout, Ajit S. Puri, Nazli Janjua, Aniel Majjhoo, Aamir Badruddin, Randall C. Edgell, Rakesh Khatri, Larry Morgan, Anmar Razak, Alicia Zha, Priyank Khandelwal, Nils H. Mueller-Kronast, Dennis J. Rivet, Thomas Wolfe, Brian Snelling, Ali Sultan-Qurraie, Shao-Pow Lin, Rajkamal Khangura, Alejandro Spiotta, Parita Bhuva, Sergio Salazar-Marioni, Eugene Lin, Abdul R. Tarabishy, Edgar A. Samaniego, Murali Kolikonda, Mouhammad A. Jumaa, Vivek K. Reddy, Pankaj Sharma, Olvert A. Berkhemer, Pieter Jan van Doormaal, Adriaan C.G.M van Es, Wim H. van Zwam, Bart J. Emmer, Ludo Beenen, Nancy Buderer, Michelle A. Detry, Anna Bosse, Todd L. Graves, Christina Saunders, Lucas Elijovich, Ashutosh Jadhav, Scott Brown, Thanh N. Nguyen, Daryl Gress, Mary Patterson, Hannah Slight, Kristine Below, Diederik W.J. Dippel, Wade S. Smith, TESLA Investigators[:: SSRN [Internet]. [cited 2024 Mar 4]. Available from: https://papers.ssrn.com/sol3/papers.cfm?abstract_id=4587818

9. Bendszus M, Fiehler J, Subtil F, Bonekamp S, Aamodt AH, Fuentes B, et al. Endovascular thrombectomy for acute ischaemic stroke with established large infarct: multicentre, open-label, randomised trial. The Lancet. 2023 Nov;402(10414):1753–63.

10. Costalat V, Lapergue B, Albucher J, Labreuche J, Henon H, Gory B, et al. Evaluation of acute mechanical revascularization in large stroke (ASPECTSIZ 5) and large vessel occlusion within 7 h of last-seen-well: The LASTE multicenter, randomized, clinical trial protocol. Int J Stroke. 2024;19(1):114–9.

11. Wei W, Zhang J, Xie S, Fan D, Chen Y, Zhong C, et al. Endovascular therapy versus medical management for acute ischemic stroke with large infarct core: Systematic review and meta-analysis of randomized controlled trials. Clin Neurol Neurosurg. 2023 Nov;234:108007.

12. Palaiodimou L, Sarraj A, Safouris A, Magoufis G, Lemmens R, Sandset EC, et al. Endovascular treatment for large-core ischaemic stroke: a meta-analysis of randomised controlled clinical trials. J Neurol Neurosurg Psychiatry. 2023 Oct;94(10):781–5.

13. Abuelazm M, Ahmad U, Abu Suilik H, Seri A, Mahmoud A, Abdelazeem B. Endovascular Thrombectomy for Acute Stroke with a Large Ischemic Core: A Systematic Review and Meta-Analysis of Randomized Controlled Trials. Clin Neuroradiol. 2023 Sep;33(3):625–34.

14. Sarraj A, Abraham MG, Hassan AE, Blackburn S, Kasner SE, Ortega-Gutierrez S, et al. Endovascular thrombectomy plus medical care versus medical care alone for large ischaemic stroke: 1-year outcomes of the SELECT2 trial. Lancet Lond Engl. 2024 Feb 24;403(10428):731–40.

15. Thrombectomy Advantage for Large-Core Strokes Widens Over 12 Months: TESLA | tctmd.com [Internet]. [cited 2024 Mar 4]. Available from: https://www.tctmd.com/news/thrombectomy-advantage-large-core-strokes-widens-over-12-months-tesla

16. Cochrane Handbook for Systematic Reviews of Interventions | Wiley Online Books [Internet]. [cited 2023 Dec 6]. Available from: https://onlinelibrary.wiley.com/doi/book/10.1002/9781119536604

17. Page MJ, McKenzie JE, Bossuyt PM, Boutron I, Hoffmann TC, Mulrow CD, et al. The PRISMA 2020 statement: an updated guideline for reporting systematic reviews. BMJ. 2021 Mar 29;372:n71.

18. The Cochrane Collaboration’s tool for assessing risk of bias in randomised trials | The BMJ [Internet]. [cited 2023 Sep 16]. Available from: https://www.bmj.com/content/343/bmj.d5928

19. Powers WJ, Rabinstein AA, Ackerson T, Adeoye OM, Bambakidis NC, Becker K, et al. Guidelines for the early management of patients with acute ischemic stroke: 2019 update to the 2018 guidelines for the early management of acute ischemic stroke: a guideline for healthcare professionals from the American Heart Association/American Stroke Association. Stroke. 2019;50(12):e344–418.

20. Almekhlafi MA, Hill MD, Roos YM, Campbell BC, Muir KW, Demchuk AM, et al. Stroke laterality did not modify outcomes in the HERMES meta-analysis of individual patient data of 7 trials. Stroke. 2019;50(8):2118–24.

21. Abuelazm M, Ahmad U, Abu Suilik H, Seri A, Mahmoud A, Abdelazeem B. Endovascular Thrombectomy for Acute Stroke with a Large Ischemic Core: A Systematic Review and Meta-Analysis of Randomized Controlled Trials. Clin Neuroradiol. 2023;33(3):625–34.

22. Kobeissi H, Adusumilli G, Ghozy S, Kadirvel R, Brinjikji W, Albers GW, et al. Endovascular thrombectomy for ischemic stroke with large core volume: An updated, post-TESLA systematic review and meta-analysis of the randomized trials. Interv Neuroradiol. 2023;15910199231185738.

23. Li Q, Abdalkader M, Siegler JE, Yaghi S, Sarraj A, Campbell BC, et al. Mechanical Thrombectomy for Large Ischemic Stroke: A Systematic Review and Meta-Analysis. Neurology. 2023;

24. Goyal M, Ospel JM, Menon B, Almekhlafi M, Jayaraman M, Fiehler J, et al. Challenging the Ischemic Core Concept in Acute Ischemic Stroke Imaging. Stroke. 2020 Oct;51(10):3147–55.

25. Broocks G, Kemmling A, Kniep H, Meyer L, Faizy TD, Hanning U, et al. Edema Reduction versus Penumbra Salvage: Investigating Treatment Effects of Mechanical Thrombectomy in Ischemic Stroke. Ann Neurol. 2023 Sep 19;

