## Supplementary file for "Endovascular thrombectomy for acute ischemic stroke with a large infarct area: an updated systematic review and meta-analysis of randomized controlled trials"

This supplemental material has been provided by the authors to give readers additional information about their work.

**Supplementary Table 1: Search Strategy for MEDLINE**

| **Number** | **Search Terms** |
| --- | --- |
| #1 | Mechanical thrombectomy [All Fields] |
| #2 | Endovascular* [All Fields] |
| #3 | “Thrombectomy” [mh] |
| #4 | #1 OR #2 OR #3 |
| #5 | Ischemic core [All Fields] |
| #6 | Large infarct [All Fields] |
| #7 | Low ASPECTS [All Fields] |
| #8 | Large baseline infarct [All Fields] |
| #9 | Large core* [All Fields] |
| #10 | Core volume [All Fields] |
| #11 | Imaging lesions [All Fields] |
| #12 | Alberta Stroke Program Early Computed Tomography Score [All Fields] |
| #13 | Massive cerebral infarction [All Fields] |
| #14 | Large hemispheric infarction [mh] |
| #15 | #5 OR #6 OR #7 OR #8 OR #9 OR #10 OR #11 OR #12 OR #13 OR #14 |
| #16 | #4 AND #15 |

**Supplementary Figure 1. Forest plot of functional independence (mRS ≤ 2) at 1 year follow-up**

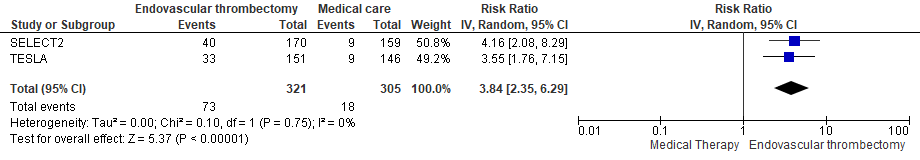

**Supplementary Figure 2. Forest plot of independent ambulation (mRS ≤ 3) at 1 year follow-up**

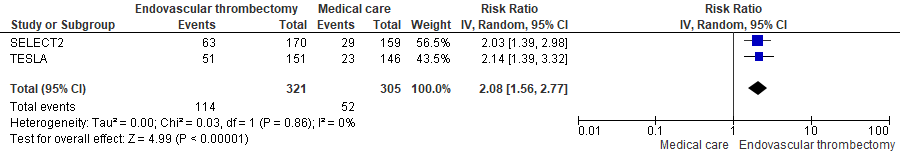

**Supplementary Figure 3. Forest plot of early neurological improvement**

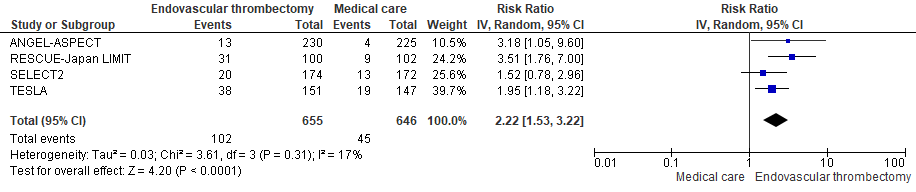

**Supplementary Figure 4. Forest plot of excellent neurological recovery (mRS ≤ 1)**

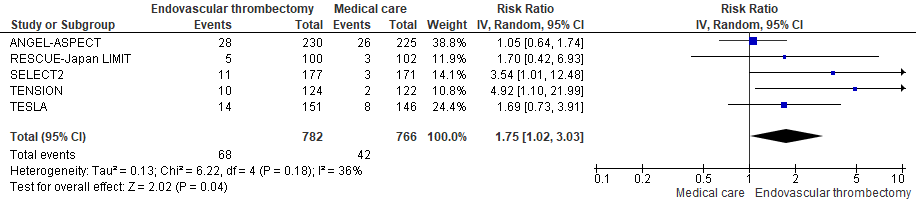

**Supplementary Figure 5. Forest plot of poor neurological recovery (mRS 4-6)**

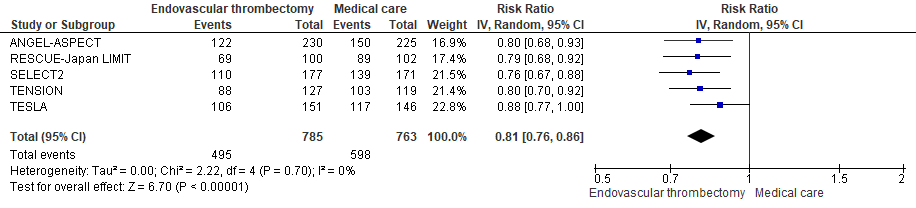

**Supplementary Figure 6. Forest plot of all-cause mortality**

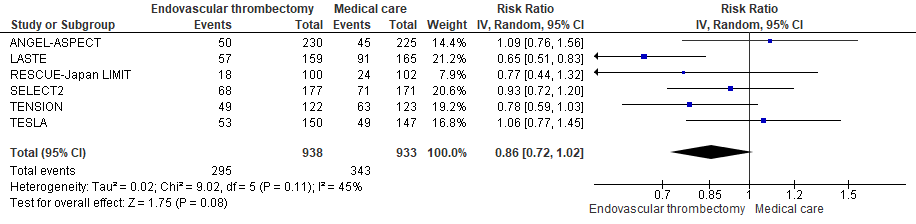

**Supplementary Figure 7. Forest plot of symptomatic intracranial hemorrhage (sICH)**

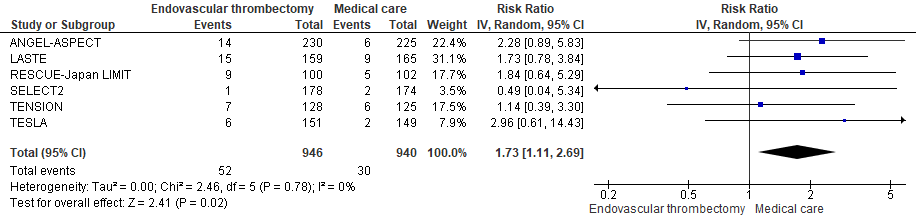

**Supplementary Figure 8. Forest plot of decompressive craniectomy**

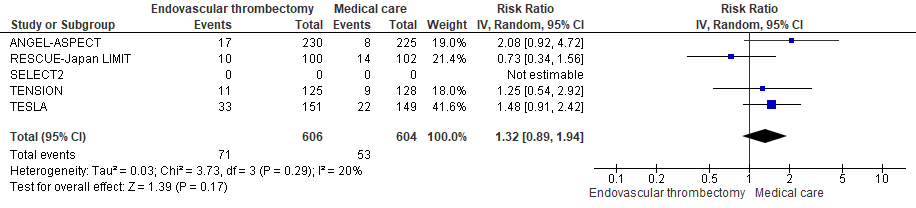

**Supplementary Figure 9. Forest plot of any intracranial hemorrhage (ICH)**

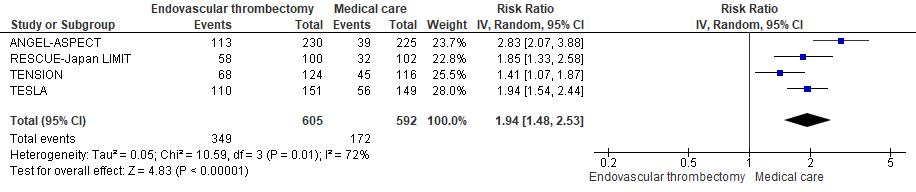

**Supplementary Figure 10. Forest plot of >1 SAE (serious adverse event)**

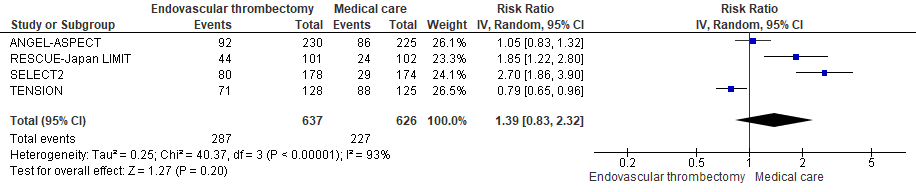
